# The relationship between patient characteristics and discharge against medical advice in patients with traumatic brain injury

**DOI:** 10.1101/2025.04.08.25324966

**Authors:** Shameeke Taylor, Tirth Patel

## Abstract

**Background:** Discharge against medical advice (DAMA), affecting up to 2% of hospital discharges, is a critical public health concern. Patients with traumatic injuries, particularly traumatic brain injury (TBI), exhibit higher DAMA rates. However, TBI-specific DAMA remains understudied. This study aims to quantify the burden of DAMA in an urban TBI cohort, analyze demographic and injury differences between patients who DAMA and those with standard discharges, and determine potential predictors of DAMA in TBI.

**Methods:** A retrospective review of TBI patients treated at an urban trauma center between 2017 and 2022 was conducted using data from our institution’s trauma registry, a subset of the National Trauma Registry of the American College of Surgeons (NTRACS). Discharge against medical advice was defined as any discharge against medical recommendations during the index hospital stay following TBI. Discharge against medical advice status was classified based on the recorded registry discharge disposition and dichotomized as DAMA and n-DAMA (non-DAMA discharge included discharge to home, inpatient facility or hospital transfer based on medical recommendation). Descriptive statistics, univariate analyses, and multivariate modeling were used to compare the DAMA and n-DAMA groups.

**Results:** This study identified 47 (3.7%) patients with DAMA status and 1214 (96.3%) without a premature discharge (n-DAMA). Younger age, male sex, Black race, shorter hospital lengths of stay, alcohol use and intentional injury were associated with DAMA in patients with TBI. Patients with n-DAMA status were more likely to be older and have Medicare insurance. No association was found between discharge against medical advice and TBI severity, GCS score, injury severity score, ventilator usage, intensive care unit days or in-hospital complications. Multivariate analysis found that intentional injury, male sex and alcohol use near the time of injury were predictive of discharge against medical advice.

**Conclusion:** TBI patients leaving against medical advice were disproportionately younger males, often with injuries linked to alcohol and violence. Given the multiple variables associated with DAMA, targeted prevention programs are crucial for this vulnerable population. Further research is necessary to understand the long-term consequences and re-injury risks after DAMA.

## Introduction

Medical providers often face situations where patients choose to leave their care before the treating physician recommends discharge, during the evaluation or treatment process. This is known as discharge against medical advice (DAMA), which is a significant public health issue, occurring in up to 2% of all hospital discharges. (1) In trauma patients, who are particularly vulnerable and at high risk, studies show that DAMA rates can range from 1.3% to as high as 7%. (2,3) The decision to discharge against medical advice can be frustrating for healthcare providers and presents serious health risks for the patient. Leaving the hospital against medical advice after a traumatic injury has been linked to higher rates of readmission within 30 days, as well as a greater likelihood of multiple readmissions following the initial discharge. (4)

Recent studies have highlighted the issue of discharge against medical advice (DAMA) among patients with traumatic brain injury (TBI). One study identified TBI as the most common diagnosis among patients who left the hospital against medical advice. (5) Kim et al. found that 2.8% of TBI patients were discharged against medical advice, with factors such as younger age, substance abuse, and self-inflicted injuries being associated with unplanned discharge. (6) Similarly, Jumah et al. reported that 2.6% of concussion patients left against medical advice, with a higher burden of comorbidities, falls, and male sex being linked to DAMA, while higher socioeconomic status was negatively correlated. (7) DAMA is also a concern for TBI patients requiring neurosurgical intervention. Jo et al. found that about 22% of TBI patients who were candidates for surgery refused the recommended procedure and left against medical advice. (8) This refusal was associated with the use of anticoagulants and the initial Glasgow Coma Scale (GCS) score. (8)

The consequences of discharge against medical advice (DAMA) for patients with traumatic brain injury (TBI) can extend well beyond the initial early discharge. When discharge is planned, providers can educate patients, prescribe necessary medications, and arrange follow-up care with community resources, as well as specialty and primary care physicians, all of which are crucial for patients recovering from TBI in the community. Disrupting this process can impede access to vital information on symptom management and follow-up care. This is particularly concerning given that at least a third of TBI patients still experience moderate to severe post-concussive symptoms three months after discharge. (9) Furthermore, between 13% and 61% of patients may not return to work within a year of the injury, depending on the severity of the injury, symptom progression and other factors. (10,11) Edwards et al. found that among patients who left against medical advice (DAMA), only 24.4% had prescribed medications documented in their charts, and only 31.3% had follow-up plans recorded. (12) DAMA not only disrupts symptom management but also significantly reduces the chances that these patients will receive the necessary follow-up care to effectively manage their condition and return to baseline functioning, which is particularly critical for those with TBI.

Identifying patients at risk for DAMA and addressing their needs during hospitalization is crucial for preventing poor outcomes. However, there is limited research on DAMA in patients with TBI, particularly within urban healthcare systems. This study aims to analyze patients with TBI who leave against medical advice (AMA) from an urban trauma center. The specific objectives are to (1) determine the number and characteristics of individuals who leave our urban hospital system after TBI against medical advice; (2) compare the demographic and injury characteristics of patients in the DAMA group versus those with an anticipated discharge; and (3) assess potential predictors of DAMA in individuals with TBI.

## Methods

This was a retrospective study using the 2017-2022 Mount Sinai Hospital trauma registry. This trauma registry was previously collected as part of the National Trauma Data Bank (NTDB). The NTDB, managed by the American College of Surgeons, is the largest voluntarily reported trauma registry in the United States. This database contains clinical and non-clinical variables, including patient demographics, diagnoses and procedures performed and hospital metrics such as patient length of stay. Because of the de-identified nature of the data used from the trauma registry, this study was deemed exempt from the institutional review board at the Mount Sinai Health System.

The Abbreviated Injury Scale (AIS) score was used to query the trauma registry for all trauma evaluations with noted traumatic head injuries. The severity of TBI ranged from mild/concussion to severe and was determined based on the Glasgow coma scale (GCS) score. TBI severity was classified as mild, moderate, or severe based on GCS scores of 13-15, 9-12, and 3-8, respectively, as recorded upon patient arrival at the emergency department. Patients who were treated primarily at our trauma center’s emergency department (ED) or transferred from an outside hospital ED to our ED for secondary trauma evaluation and management were included. Patients with penetrating (gunshot and stab wounds) and blunt trauma (motor vehicle collisions, falls, blunt injury from assaults etc.) were considered for this study. Only those who survived to emergency department trauma evaluation (time of death not called in the field or at time of ED arrival) were included. Discharge against medical advice status was classified based on the recorded registry discharge disposition and dichotomized as DAMA and n-DAMA (non-DAMA discharge which included discharge to home, inpatient facility or transfer). DAMA classification was for any discharge against medical advice from the evaluating trauma hospital (ED or inpatient) during the index hospital visit after suffering a TBI.

This study examined multiple patient demographic and injury characteristics, including age, sex, race/ethnicity, TBI severity, Glasgow Coma Scale (GCS) score, Injury Severity Score (ISS), alcohol use, orthopedic injuries, complication burden, insurance type, injury intent and hospital discharge outcome. Additionally, we assessed the length of mechanical ventilation, ICU stay, and overall hospital stay. The presence of orthopedic surgeon consultations and documented traumatic fractures using the AIS diagnosis code were used to identify orthopedic injuries. Alcohol use near the time of injury was confirmed through positive blood alcohol tests or documented clinical suspicion by the treating physicians. Minority status was determined by patient racial and ethnic self-classification as other than non-Hispanic White. The complication burden was calculated as a composite score, based on the total number of complications experienced by each patient. The complications based on NTDB classification include cardiac arrest, unplanned return to the operating room, unplanned intubation, unplanned extubation, inpatient alcohol withdrawal, ventilator-associated pneumonia, and acute respiratory distress syndrome. The designation of trauma due to violence or acts of self harm in the database determined injury intent or intentionality. Intentional trauma types included assault, gun or stab wounds and suicide attempts.

We began by conducting descriptive analyses of the overall TBI cohort, as well as the DAMA and n-DAMA subgroups. Next, we compared patient characteristics based on the presence or absence of DAMA status. We applied the Chi-squared or Fisher’s exact test for categorical variables, while t-tests or Wilcoxon rank sum tests were used for continuous variables. To identify factors associated with DAMA status, multivariate logistic regression was employed. The results of the regression are presented as adjusted odds ratios with 95% confidence intervals (95% CIs). A p-value of less than 0.05 (two-sided) was considered statistically significant. Multicollinearity was assessed using the variance inflation factor (VIF), and no significant evidence of collinearity was found to affect our model’s results. Model calibration was evaluated with the Hosmer-Lemeshow statistic, and discrimination was assessed by calculating the area under the receiver operating characteristic curve. All analyses were performed using SAS On Demand for Academics (Cary, NC, USA).

## Results

Based on the inclusion and exclusion criteria for this study, 1261 patients with TBI were included in the analysis. Of the total cohort, 47 patients (3.7%) were discharged against medical advice (DAMA) and 1214 patients did not have a premature discharge (n-DAMA) from the hospital after TBI. Table 1 describes the demographic and injury related characteristics for the cohort. The majority of patients were male sex accounting for approximately 65% of the sample. Black and Hispanic TBI patients comprised approximately 39.7% of participants. Most patients sustained a mild TBI with this group representing 85% of the group, while moderate and severe TBI accounted for 5.3% and 9.2% of the sample respectively. Medicare and commercial insurance were the most commonly held types at 35.1% and 29.4% respectively.

**Table 1:** TBI Cohort characteristics for DAMA study.

| Variable | Mean ± SD or n (%) |
| --- | --- |
| Age | 66.4 ± 19.9 |
| Sex |  |
| Male | 820 (65%) |
| Female | 441 (35%) |
| Race/Ethnicity |  |
| White | 255 (20.2%) |
| Black | 186 (14.8%) |
| Hispanic | 314 (25.0%) |
| Asian | 36 (2.8%) |
| Other | 456 (36.1%) |
| Unknown | 14 (1.1%) |
| **Insurance** |  |
| Commercial | 371 (29.4%) |
| Medicare | 443 (35.1%) |
| Medicaid | 283 (22.5%) |
| Other | 164 (13.0%) |
| **Glasgow Coma Scale score** | 13.6 ± 3.1 |
| **Traumatic Brain Injury Severity** |  |
| Mild | 1075 (85.2%) |
| Moderate | 67 (5.3%) |
| Severe | 119 (9.5%) |
| **Injury Severity Score** | 14.1 ± 7.8 |
| **Orthopedic injuries** | 203 (16.1%) |
| **Complications** | 90 (7.1%) |
| **In-Hospital Death** | 100 (7.9%) |
| **Total Length of Stay** | 7.9 ± 15.7 |

Table 2 compares the characteristics of DAMA and n-DAMA patients. DAMA patients were younger (56.1 vs 66.6 years; <.0001) and had a higher percentage of males (85.1% vs 64.3%; p=0.0033) and individuals identifying as Black (25.5% vs 14.3%; p=0.0336) compared to n-DAMA patients. DAMA patients also had a higher rate of alcohol use near the time of injury (42.6% vs 18.1%; <.0001) and were more likely to present to the ED with intentional injuries (27.7% vs 8.6%; p<.0001). Conversely, DAMA patients had a lower proportion of Medicare insurance (10.6% vs 36.1%; 0.0001), identification as female (14.9% vs 35.7%; p=0.0033), and shorter lengths of stay in the hospital for their initial head injury visit (3.1 vs 8.1 days; <.0001). No significant differences were observed between the DAMA and n-DAMA groups in terms of ICU days, ventilator use, intubation, race or ethnicity (non-Black), overall minority status, TBI severity, Medicaid or commercial insurance, presence of orthopedic injuries, complication burden, ISS, or GCS at arrival.

**Table 2:** Comparison of n-DAMA and DAMA status.

| **Variable** | **DAMA** | **n-DAMA** | **Significance (p)** |
| --- | --- | --- | --- |
| Age | 56.1±18.3 | 66.6±19.9 | <0.0001 |
| Male, n (%) | 40 (85.1) | 780 (64.3) | 0.0033 |
| Female, n (%) | 7 (14.9) | 434 (35.8) | 0.0033 |
| Black race, n (%) | 12 (25.5) | 174 (14.3) | 0.0336 |
| White race, n (%) | 7 (14.9) | 248 (20.4) | 0.3540 |
| Asian race, n (%) | 2 (4.3) | 34 (2.8) | 0.5568 |
| Hispanic ethn., n (%) | 10 (21.3) | 304 (25.0) | 0.5582 |
| Length of stay (mean±SD) | 3.1± 4.6 | 8.1± 16.2 | <0.0001 |
| ISS (mean± SD) | 11.7± 4.6 | 14.0± 7.9 | 0.1460 |
| GCS score on arrival (mean± SD) | 14.0± 2.6 | 13.6± 3.1 | 0.5091 |
| ICU days (mean± SD) | 0.9± 1.7 | 2.3 ± 6.4 | 0.2459 |
| Ventilator days (mean± SD) | 0.3± 1.0 | 1.0 ± 3.8 | 0.1444 |
| Alcohol present, n (%) | 20 (42.6) | 216 (18.1) | <0.0001 |
| Orthopedic injury, n (%) | 3 (6.4) | 200 (16.5) | 0.0648 |
| Complication burden (mean± SD) | 0.04 (0.2) | 0.20 (0.5) | 0.1609 |
| Intentional injury, n (%) | 13 (27.7) | 104 (8.6) | <.0001 |
| **Insurance Type** |  |  |  |
| Commercial, n (%) | 13 (27.7) | 358 (29.5) | 0.7871 |
| Medicare, n (%) | 5 (10.6) | 438 (36.1) | 0.0003 |
| Medicaid, n (%) | 16 (34.0) | 267 (22.0) | 0.0521 |
| **TBI severity** |  |  |  |
| Mild, n (%) | 42 (89.4) | 1033 (85.1) | 0.4178 |
| Moderate, n (%) | 3 (6.4) | 64 (5.3) | 0.7460 |
| Severe, n (%) | 2 (4.3) | 117 (9.6) | 0.2156 |

Abbreviations: GCS, glasgow coma scale; ICU, intensive care unit; ISS, injury severity score; TBI, traumatic brain injury.

Regression modeling for the outcome of DAMA demonstrated that the intentionality of injury, male sex and alcohol use predicted DAMA status. These data are depicted in Table 3. Intentional injury was associated with 2.615x greater odds of DAMA than unintentional injury. Male sex was associated with 2.451x greater odds of DAMA than female sex. Alcohol use was associated with 2.378x greater odds of DAMA than patients without a positive alcohol test or clinical suspicion of intoxication. ISS did not produce statistically significant associations with DAMA status. This model had moderate discrimination with an AUC of 7.221 and good calibration based on goodness of fit testing.

**Table 3:** Regression model analysis for DAMA status.

| Effect | Point Estimate | 95% Confidence Limits | Pr > ChiSq |
| --- | --- | --- | --- |
| Intentional | 2.615 | (1.286-5.315) | 0.0079 |
| Male Sex | 2.451 | (1.057-5.684) | 0.0367 |
| ISS | 0.960 | (0.917-1.006) | 0.0869 |
| Alcohol use | 2.378 | (1.272-4.447) | 0.0067 |

Abbreviations: ISS, injury severity score.

## Discussion

Traumatic brain injury (TBI) is a significant global health concern, affecting approximately 1.7 million individuals in the United States annually and disproportionately impacting young people. (13) Despite advancements in TBI treatment, a concerning number of patients choose to leave the hospital against medical advice (DAMA), interrupting their care and potentially compromising their recovery.

In our single-center study we explored the factors associated with DAMA among TBI patients. Our analysis revealed a 3.7% DAMA rate, consistent with prior research. (1,2,3) Our regression analysis revealed that intentional injury, male sex and alcohol use were independent predictors of DAMA highlighting the complex interplay of demographic, clinical, and social factors influencing this decision.

Our findings corroborate previous studies demonstrating that TBI patients who choose DAMA often differ significantly from those who complete their treatment. Specifically, DAMA patients were more likely to be young, male, and Black, mirroring demographic profiles observed in general trauma populations. (4,5) This suggests that young Black men may be particularly vulnerable to factors contributing to unplanned discharge, such as distrust of the medical system, or socioeconomic barriers. However, the increased risks of DAMA in this racial group are possibly confounded by socioeconomic status. In previous literature evaluating the risk of DAMA in Black patients found that after adjusting for possible confounders including socioeconomic status there was no significant difference in DAMA between Black patients and their White counterparts. (14) Future studies should focus on investigating discharge patterns to better understand racial/ethnic differences and potential mediators of increased risk in order to develop strategies to mitigate potential unanticipated discharge.

Substance use, particularly alcohol, is also strongly associated with DAMA. (15) Alcohol is known not only be associated with traumatic injury (16) but also the risk of premature departure. (15) Therefore, alcohol use can impact both primary injury prevention and the secondary prevention of injury.(17) These patients may experience a confluence of medical, psychiatric, and social challenges that contribute to their premature departure. Acute alcohol intoxication may also lead to impulsive behavior and lack of proper self-determination skills with concomitant risk taking behaviors and lack of awareness in the severity of their injuries or their own capacity to understand their current medical needs.(17) These concerns may lead to significant traumatic injuries and early departure from the healthcare facility prior to clinician recommended disposition where they may be at continued risk of injury upon discharge as they may continue the same behaviors or suffer from the same untreated medical conditions that increased their risk of injury. This underscores the need for integrated care models addressing substance abuse, mental health, and social support alongside medical treatment. Comprehensive care extending beyond the immediate physical injury may better support these vulnerable patients and reduce DAMA incidence.

Our study found that injury intent was associated with DAMA. Intentional trauma was associated with DAMA when compared to trauma from unintentional injuries such as falls and motor vehicle accidents. We postulate that this increased risk is due to an association with multiple potential factors. Intentional injury is due to violence which includes assaults and self-inflicted trauma (suicide attempts). Violent injuries are more likely in individuals with low socioeconomic status, neighborhood poverty and in individuals with mental health and substance abuse concerns. (18-20) These factors are also associated with an increased risk of DAMA and may mediate the risk of unanticipated discharge with intentional injuries.(15, 21, 22) A better understanding of the intersectional relationships of these social determinants is warranted to reduce potential DAMA in at risk patients.

While our study did not find a significant association between TBI severity and DAMA, which contrasts with some prior research, this may be attributable to our specific sample characteristics or the method of TBI severity assessment. Nevertheless, the potential for long-term impairments even in mild TBI cases makes any disruption of care concerning. Our study found significantly shorter length of stay among DAMA patients, highlighting the disruption of care and potential loss of opportunities for patient education, medication reconciliation, and follow-up planning. As Edwards et al. note, DAMA patients are less likely to have documented medication prescriptions or follow-up plans, affecting their recovery. (6) This is particularly concerning for TBI patients given the prevalence of post-concussive symptoms and the need for patient education and ongoing support.

This study is limited by its retrospective, single-center design, which may impact generalizability. Furthermore, while the NTDB is a large database, it may not fully capture the complex social determinants influencing DAMA. Future research, including qualitative studies or linkage with socioeconomic datasets, could explore these factors further. Despite these limitations, this study provides valuable insights into DAMA in TBI patients, emphasizing the need for interventions addressing the complex needs of this vulnerable population.

### Future Directions

Future research should focus on developing and evaluating interventions to reduce DAMA among TBI patients. These interventions should be tailored to address the specific needs of high-risk individuals, such as those with substance abuse issues or mental health problems. Additionally, future studies should examine the long-term outcomes of DAMA patients, including rates of readmission, disability, and mortality. Qualitative research methods may also be useful to explore the reasons why patients choose to leave against medical advice and to identify barriers to adherence to medical recommendations.

## Conclusion

In conclusion, this study provides valuable insights into the characteristics and predictors of DAMA among TBI patients in an urban trauma center. Our findings highlight the importance of addressing modifiable risk factors, such as alcohol use, to reduce the incidence of DAMA and improve outcomes for this vulnerable population. By implementing targeted interventions and improving discharge planning, healthcare providers can help to ensure that TBI patients receive the care and support they need to recover from their injuries.

## Data Availability

All data in the present study are available upon reasonable request to the Mount Sinai Health System, Department of Surgery and completion of an IRB application for use of the data.

